# A novel biomechanical injury risk score demonstrates correlation with lower limb posterior chain injury in fifty elite level rugby union athletes

**DOI:** 10.1101/2021.02.02.21250855

**Authors:** Rhys Hughes, Matt Cross, Keith Stokes

## Abstract

**Objectives:** Lower limb posterior chain injury (PCI) is common amongst athletic populations, with multi-factorial risk factors including age, previous injury, strength measurements, range of motion and training load. Biomechanics are commonly considered in the prevention and rehabilitation of PCI by performance staff. However, there is no documented testing method to assess for associations between biomechanics and PCI. The aim of this study was to investigate whether there is an association between an easily applicable, novel biomechanical assessment tool and PCI.

**Methods:** Fifty male elite-level rugby union athletes (age 22.83±5.08) participating in the highest tier of England were tested at the start of the 2019 pre-season period and PCIs (N=48) were recorded over the 2019/20 playing season. Participants’ biomechanics were analysed using two-dimensional video analysis against an Injury Risk Score (IRS) system in the performance of the combined movement – prone hip extension and knee flexion. Participants’ biomechanics in carrying out this movement were scored against the 10-point IRS, where the more compensatory movement recorded sees an increase in an individual’s IRS. Participants’ IRS were then compared against the number of PCI sustained and Spearman’s correlation coefficient was utilised for analysis.

**Results:** There is a good significant association between IRS and PCI (R=0.573, p<0.001). Linear Regression demonstrated that an increase of 1 in IRS was associated with a 35% increase in PCI incidence (R^2^=0.346).

**Conclusion:** A good significance between the IRS and PCI provides preliminary support for its use as an injury risk assessment tool.

## INTRODUCTION

The term posterior chain injury (PCI) is commonly used in relation to injuries to the posterior musculoskeletal system of the lower limb.[1-2] PCI is commonly observed within athletic populations, with a reported time loss of 10 hours for every 1000 playing hours within elite level sport.[3] The most prevalent PCIs involve structures within the posterior musculoskeletal system such as the trunk, pelvis, hamstring, and calf complexes.[4-8] This has subsequently led to a large amount of research being conducted that focuses on these areas of PCI, particularly during high velocity activities such as sprinting. Sprinting requires multiple structures of the posterior musculoskeletal system to work concurrently.[8-9] Occurrence of PCI is attributed to a failure and a loss of concurrence somewhere within this system.[9-10] Therefore, it is no surprise that PCI prevalence is high in many sports that require sprinting, including rugby union,[3] soccer,[11] American football [12] and athletics [13]. Of the structures within the posterior musculoskeletal system, the hamstring complex is most frequently affected.[14]

Research in athletic populations has found the non-modifiable risk factors of: 1) greater age and 2) previous injury to be predictors of PCI.[15-16] Modifiable risk factors suggested to influence PCI include deficits in strength, particularly eccentric strength in hamstring injury;[17] overload in training volume;[18-19] sprint performance;[20] and reduced ranges of motion.[21] The most influential finding regarding modifiable risk factors relating to PCI management has arguably been the introduction of the Nordic hamstring extension (NHE) in 2001.[4, 17, 22-24] Many exercise programmes focusing on the prevention or rehabilitation of PCI aim to influence these modifiable factors and include some form of coached lower limb biomechanical positioning during exercise prescription.[25] However, there is a lack of evidence of the effect biomechanics may have on PCI specifically. Whilst pelvic and lower limb positioning is assessed in many ways by practitioners in sport performance-medical settings, until recently [20, 26-27] there has been no evidence of a relationship between biomechanics and PCI. It is also worth noting, there is currently no uniform clinical assessment method of pelvic or lower limb mechanics that relates to PCI.

A multi-modal intervention of bed-based treatment, mobility and exercise therapy has demonstrated positive influence on pelvic biomechanics when assessed using an inertial sensor.[26] The authors suggest a neutral pelvis position and relative limb alignment could reduce hamstring strain injury risk, however there is a lack of uniform assessment and comparison to injury incidences to conclude this. Further research,[20] also supports a link between these factors highlighting athletes who have sustained a PCI present with an increased amount of anterior pelvic tilt through the gait cycle of running when compared against those who do not. Yet, no conclusion was drawn as to whether this was the cause or effect of the injuries and no evidential comparison can be made. The role of muscle co-contraction in biomechanics when analysing electromyography (EMG) patterns within soccer players has also been investigated.[27] Athletes who demonstrated an inefficient co-contraction pattern when recruiting specific stabilising muscles within the posterior musculoskeletal system (lumbar spine erectors, gluteus maximus and hamstrings) were up to 8 times more susceptible to PCI.[27] It is evident association between an individuals’ biomechanics and PCI risk may be present but need further evidential support with direction on a uniform assessment method.

This study assesses the association between an easily applicable, novel, clinician-assessed biomechanical injury risk score (IRS) and the number of PCI amongst a group of elite-level rugby union athletes.

## METHODS

### Study Participants

Fifty adult male professional rugby union athletes aged between 18 and 35 years (22.8±5.1) were recruited at the start of a full playing season (1^st^ July 2019) and were considered relatively well rested after a minimum of 5 weeks break between playing seasons. The participants were injury free and considered available for full training at the time of testing. Informed consent was gained from all participants and ethics were appropriately obtained from the Research Ethics Approval Committee for Health at the University of Bath, reference number EP18/19029.

### Study Design

This study investigates the association between a clinician assessed injury risk score (IRS) system and PCI sustained during one full playing season. PCI was defined as any non-contact injury to the foot (palmar aspect), calf complex (including the achilles tendon), hamstring muscle group, gluteal muscle group, lumbar spine, and thoracic spine regions that led to time loss in training and/or matches. PCI included soft tissue disruption categorised using the British Athletics Muscle Injury Classification, [28] which details tissue disruption of all types between grades 0-4; discopathy, neuropathy, tendonopathy and overload injuries were also included. Contact injuries from direct impact were excluded from the study as direct impact injuries are not deemed to be affected by biomechanics.

### The Injury Risk Score (IRS)

A visual representation of the testing procedure is presented in figure 1 and an example of the IRS system criteria in figure 2. This novel testing protocol is clinician assessed and it is not documented in empirical literature. However, the assessment method uses a modified approach that utilised EMG in assessing prone hip extension (PHE), which demonstrated good levels of reliability and validity.[27] The modification translated into this study sees the replacement of EMG with palpable assessment of gluteal muscle tone, the addition of prone knee flexion (PKF) and observation of biomechanics retrospectively analysed using two-dimensional video analysis. Biomechanics are assessed against the IRS system in the performance of combined PHE and PKF (figure 1), subsequently providing greater comparison to the gait cycle than PHE alone, where PHE and PKF are performed concurrently.[20] Participants can score a maximum of 10 points following the IRS system criteria – the more biomechanical change observed from the participants anatomical prone starting position, the higher the score. The 5 compensatory criteria include: 1) loss of palpable co-activation in either gluteal muscle, 2) lumbar spine extension, 3) lift of the anterior superior iliac spine from the plinth, 4) An increase in hip external rotation 5) An increase in hip abduction. If the athlete displays a compensatory movement pattern for one of the 5 criterion, 1 point would be scored per limb, left (maximum 5 points) and right (maximum 5 points) with a maximum of 10 points achievable.

**Figure 1:**
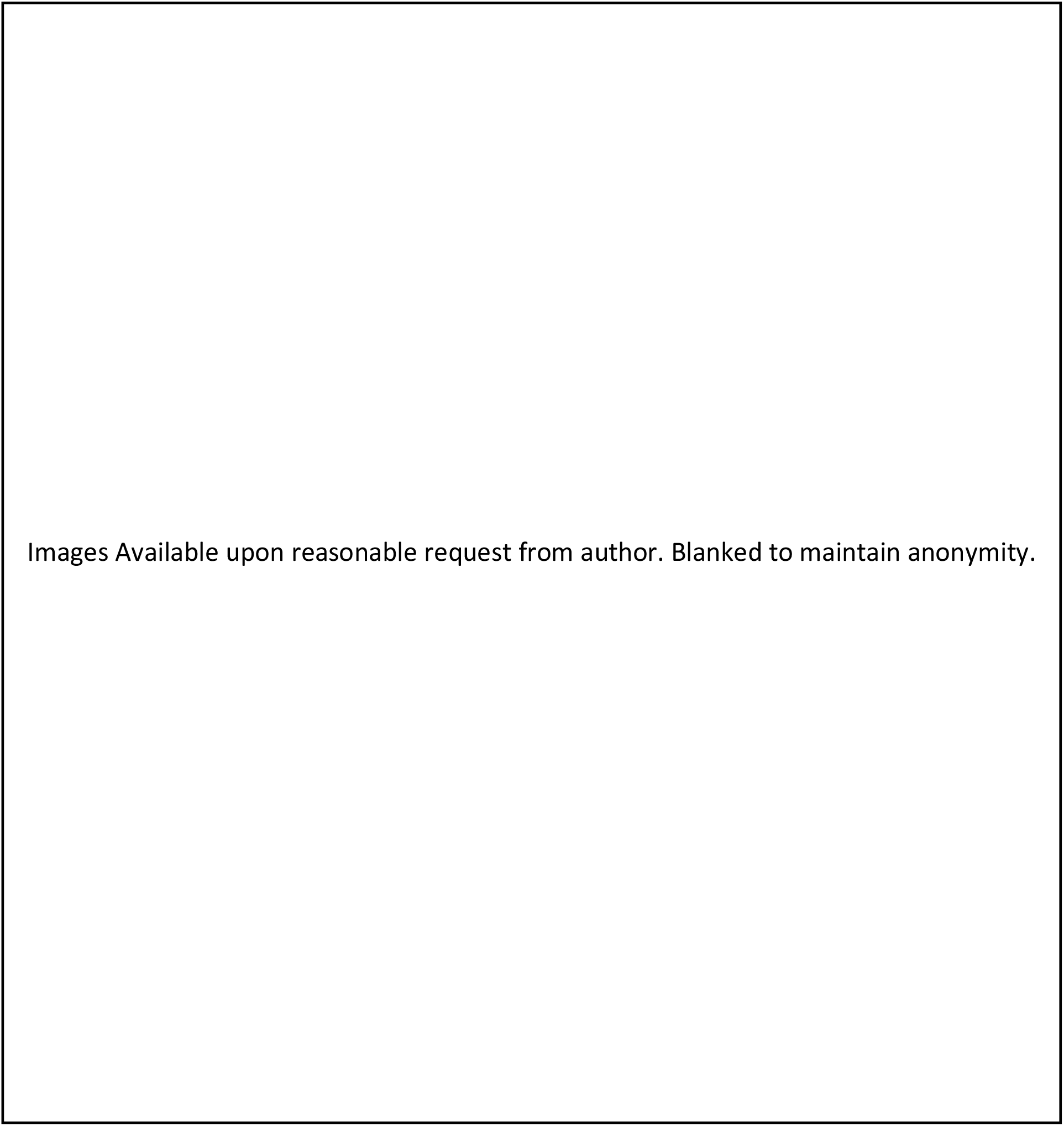
Still image views of longitudinal (left) and transverse (right) views of biomechanical deficit and limb asymmetry when using the IRS system. Starting Position (1a), Testing Left Limb (1b) and Testing Right Limb (1c).

**Figure 2:**
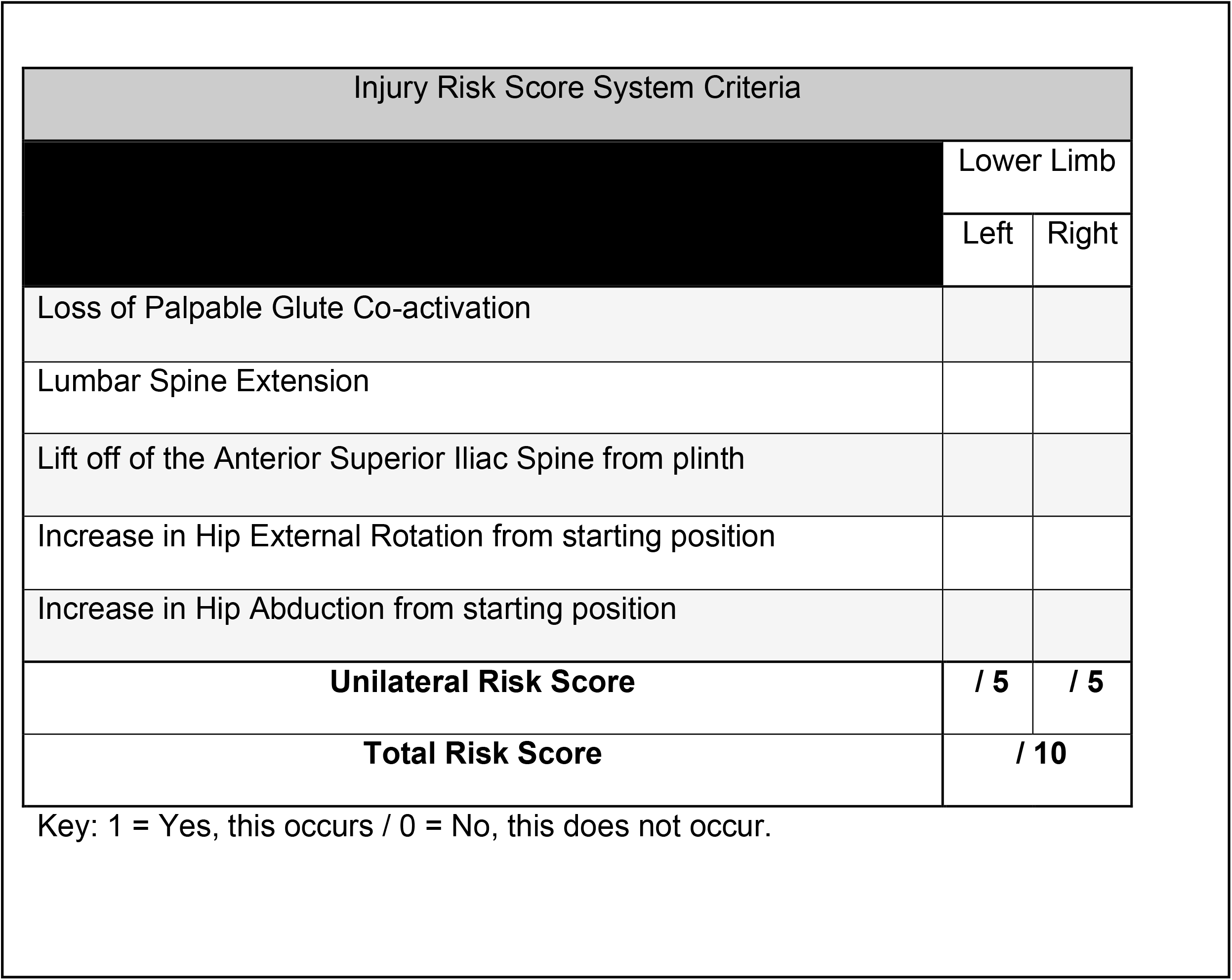
IRS System Criteria.

As the IRS system is novel, a repeatability pilot was performed prior to the data collection and results analysed using Fleiss Kappa to assess inter-rater reliability (IRR). The lead researcher educated and familiarised four assessors in the IRS system. Each clinician had a minimum of five years’ experience in musculoskeletal medicine. The clinicians were blinded and asked to analyse a small pilot group (four participants), against the IRS system. Results from this pilot study were interpreted as suggested by McHugh,[29] whereby (R=>0.90 – excellent, R=0.80-0.89 – very good, R=0.60-0.79 – good, R=0.40-0.59 – moderate and R=0.21-0.39 – minimal). Completing this pre-trial pilot demonstrated good levels of IRR (R=0.651, p <0.001) and allowed for some minor adjustments in the logistics around participant testing, reflected in the testing procedure section.

### Testing Procedure

Each participant started in a prone position on a treatment plinth with arms placed by their side or above their head (as in figure 1) to negate the use of upper limb stabilisation during testing. The testing clinician then placed their index finger and thumb of one hand on the horizontal plateau of the gluteus maximus to assess palpable gluteal activation.[30] Prior to testing, each participant was read the same script by the testing clinician, *“Tense both buttock muscles and maintain this contraction for the duration of the movement, until told the test is over. Now bend your left knee to 90 degrees and lift that knee off the bed”*. The testing practitioner would then say, *“The test is over”*. The participant then relaxes, resuming the prone starting position. The opposite side would then be tested with the same instructions where *“left knee”* was replaced with *“right knee”*. Each participant was given a 2-minute testing window and participants were tested consecutively. The testing clinician retrospectively assessed each participant through the combined PHE and PKF movement against the IRS criteria (table 1), using video analysis. The only live score recorded during the testing protocol was palpable gluteal activation as muscle tone was deemed unrecordable by video analysis. All scores were recorded on a laptop using spreadsheet software.

### Setting and Equipment

Equipment included: a password protected external hard drive; computer compatible with spreadsheet software and the Statistical Package for the Social Sciences (SPSS) version 25 [31] for analysis; height adjustable treatment plinth (plinth height was maintained throughout); three high resolution video recording cameras (Panasonic HC-V770 50x Zoom 4k Full HD) with tripods, capable of recording in two dimensions at perpendicular angles in line with the horizontal surface of the treatment plinth. Video cameras were synchronised to record simultaneously from two angles at the start of testing. Recording angles included a lateral view of both left and right sides of the participants and a caudal to cephalad view. Footage was cropped to allow for time efficiency when assessing IRS by deleting plinth transfer footage between participants using video analysis software.[32] This method of video augmentation in movement analysis is well validated [33-34] and has proven reliable when qualitatively assessing human biomechanics against quantitative criteria.[35-37]

### Data Collection

Data collection took place within the physiotherapy room of an English premiership rugby union club. Participants were required to spend a maximum of 2 minutes each (including transfer time) in a prone position on the treatment plinth. Testing for all participants took approximately 2 hours including equipment set up. Data was collected from all participants on day one of pre-season testing. The number and location of PCIs for each participant were then recorded for the duration of one full English premiership rugby playing season using digital spreadsheet software as part of departmental injury surveillance. This was standardised using the definition of PCI as outlined in the study design and the same individual was responsible for data recording to ensure continuity and avoid potential for bias.

### Outcome Measures

The two main outcome measures for this study were PCI (dependent variable) and the IRS (independent variable). The IRS was measured quantitatively from analysis of the qualitative data collected against the IRS and cross referenced against the recorded video by the lead researcher. The number of PCI sustained by each player across a 10-month playing season was captured at the end of the season.

### Statistical Analysis

Data analysis using SPSS version 25 [31] was utilised for data comparisons and assessment of correlation. Prior to statistical analysis both IRS and PCI variables were assessed for homogeneity of variance and normal distribution using the Kolmogorov-Smirov and Shapiro-Wilk tests. As assumptions of homogeneity and normal distribution were not met, a Spearman’s correlation co-efficient (SCC) was conducted where R was interpreted as (R=>0.90 – very strong, R=0.70-0.89 – strong, R=0.40-0.69 – good, R=0.10-0.39 – weak and R=0-0.09 – negligible). [38]

To reduce the likelihood of any type 1 errors within the group (N=50) a critical alpha (p) of <0.05 was utilised. Finally, a non-parametric linear regression was conducted to demonstrate whether there was a linear relationship between PCI and IRS, and if so to what degree the variables were related. To ensure methodological rigour throughout the study adheres to STROBE [39] reporting guidelines.

### Patient and Public Involvement

Participants were first involved in research during the first day of data collection when informed consent was obtained, and their IRS was recorded. The research question and outcome measures were developed by the authors, and participants were informed of these using a patient information sheet prior to data collection. Participants were not involved in study design, recruitment or conduction and they were not asked to assess the burden or time required to participate in the study. In the dissemination of the results the participants will receive their individual IRS followed by a discussion around their PCI risk with advice around reducing this risk.

## RESULTS

Fifty subjects participated in the study (age 22.83±5.08). The mean IRS score across the population was 5.80±1.74 and participants IRS ranged from 3 to 10 with a median of 7. A total of 48 PCIs were observed in 20 of the participants across the playing season, as displayed in figure 3. Twenty participants did not suffer a PCI, eighteen suffered 1 PCI, seven suffered 2 PCIs, four suffered 3 PCIs and one suffered 4 PCIs. The average number of PCIs across the population was 0.90±1.03 per participant. The most frequently observed PCIs were lumbar spine – (muscle cramp/ stiffness/ myofascial trigger points) and hamstring – biceps femoris (grade 2) both recording 7 incidences across the season (figure 3). The number of biomechanical compensations observed across the group was 320, with some common themes in compensation, as displayed in table 2.

**Table 2:** IRS system points scored for each biomechanical compensation.

|  | Loss of Palpable Glute Co-activation | Lumbar Spine Extension | Lift off of the Anterior Superior Iliac Spine from plinth | Increase in Hip External Rotation from starting position | Increase in Hip Abduction from starting position | **TOTAL** |
| --- | --- | --- | --- | --- | --- | --- |
| Total points for each biomechanical characteristic | 62 | 95 | 71 | 54 | 38 | **320** |
| Percentage (%) | 19% | 30% | 22% | 17% | 12% |  |

**Figure 3:**
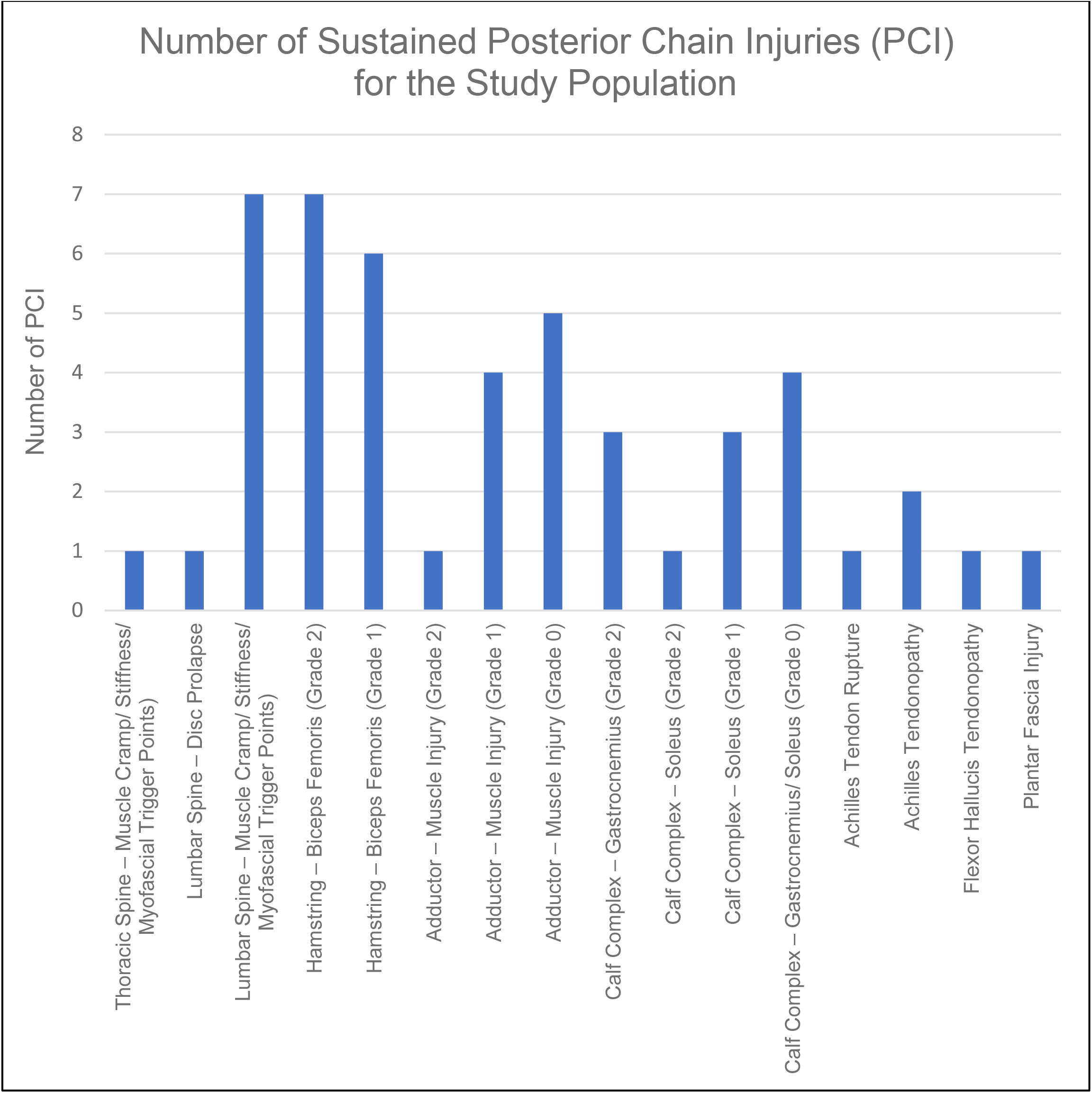
Number of Posterior Chain Injuries (PCI) for the study population.

Spearman’s correlation demonstrated a good positive association between participant IRS and number of PCIs (R=0.573 (p <0.001)) (figure 4). Linear regression identified that for every 1-point increase in IRS there was a 35% increase for a further PCI (R^2^=0.346) (figure 4).

**Figure 4:**
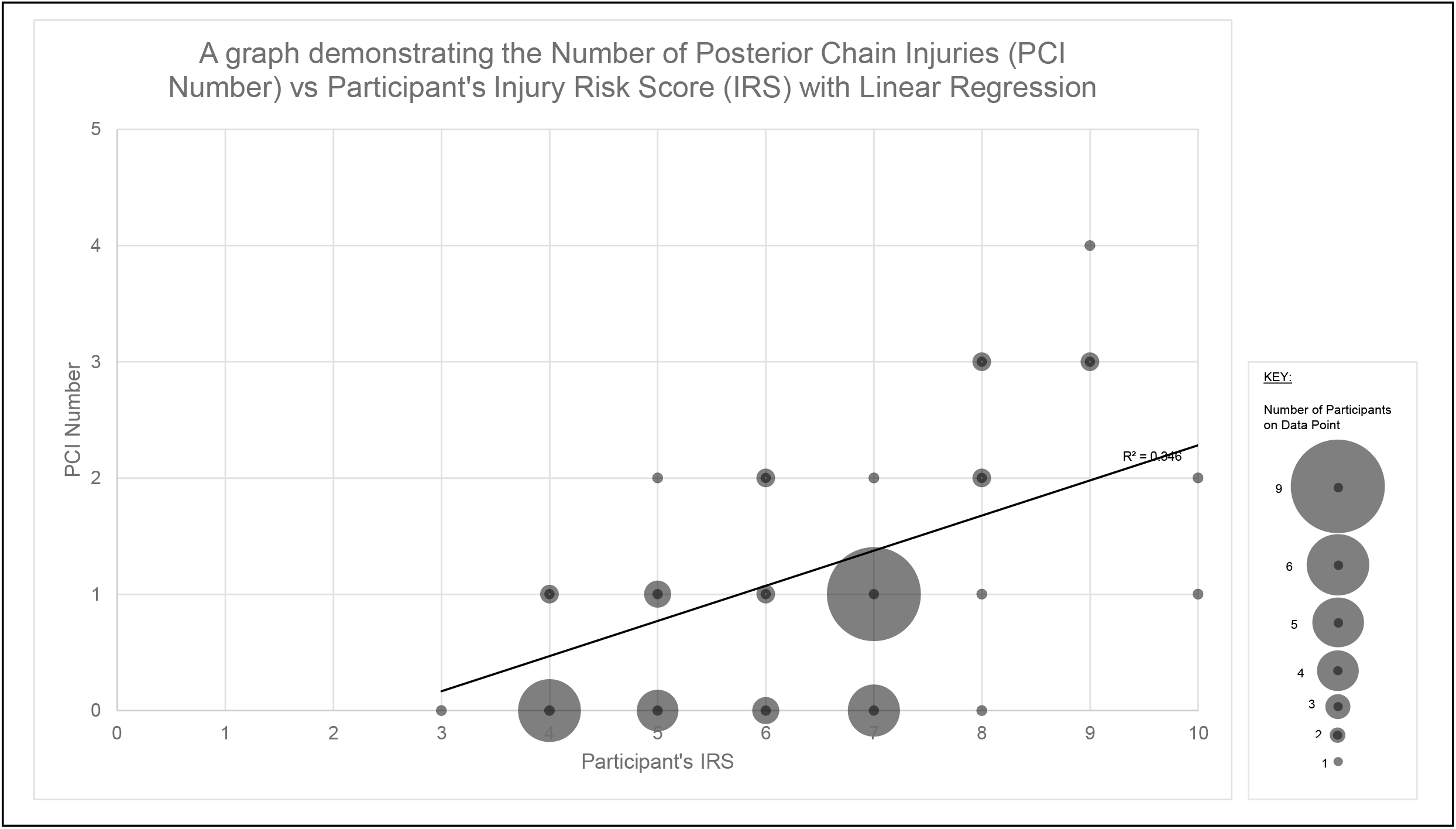
A graph demonstrating the Number of Posterior Chain Injuries (PCI Number) vs Participant’s Injury Risk Score (IRS) with Linear Regression.

## DISCUSSION

With significance demonstrating association between the IRS and PCI an increase in the identified biomechanical compensations are suggestive of increasing an individual’s risk of PCI. These results also provide confidence in the IRS as a biomechanical assessment tool for injury risk of PCI and provide guidance on biomechanical areas for intervention. Hence, practitioners may use the IRS alongside other pre-existing methods of identifying PCI risk, whilst guiding injury prevention. The IRS also has capacity to provide practitioners with areas for biomechanical improvement in the prevention of PCI when considering table 2. These suggest target areas for reducing an individual’s IRS when considering reduction of PCI risk. For example, lumbar spine extension is highlighted as the most common compensatory movement in the IRS across the population. With good levels of significance in the association between PCI and the IRS assessment tool it could be suggested that reducing lumbar spine extension which in turn lowers an individual’s IRS could reduce the PCI risk, although further investigation would be required to prove this.

It is also worth highlighting the observational differences in IRS as highlighted with the yellow markers in figure 5. The transverse views of the IRS clearly demonstrate an example of the possible biomechanical asymmetry present when performing combined prone PHE and PKF (figure 5). Figure 5 further demonstrates how observational differences may translate from the prone IRS testing position to upright running using the ALTIS kinogram analysis.[40] It is evident in the transverse views of the IRS that lower limb abduction and external rotation are increased from the midline starting position in the participants right lower limb compared to left, this is also evident at toe off in the kinogram analysis – see corresponding points A and B on figure 5. This may provide some practical explanation of how the correlation between IRS and PCI occurs during function, further promoting the use of the IRS in the management of PCI.

**Figure 5:**
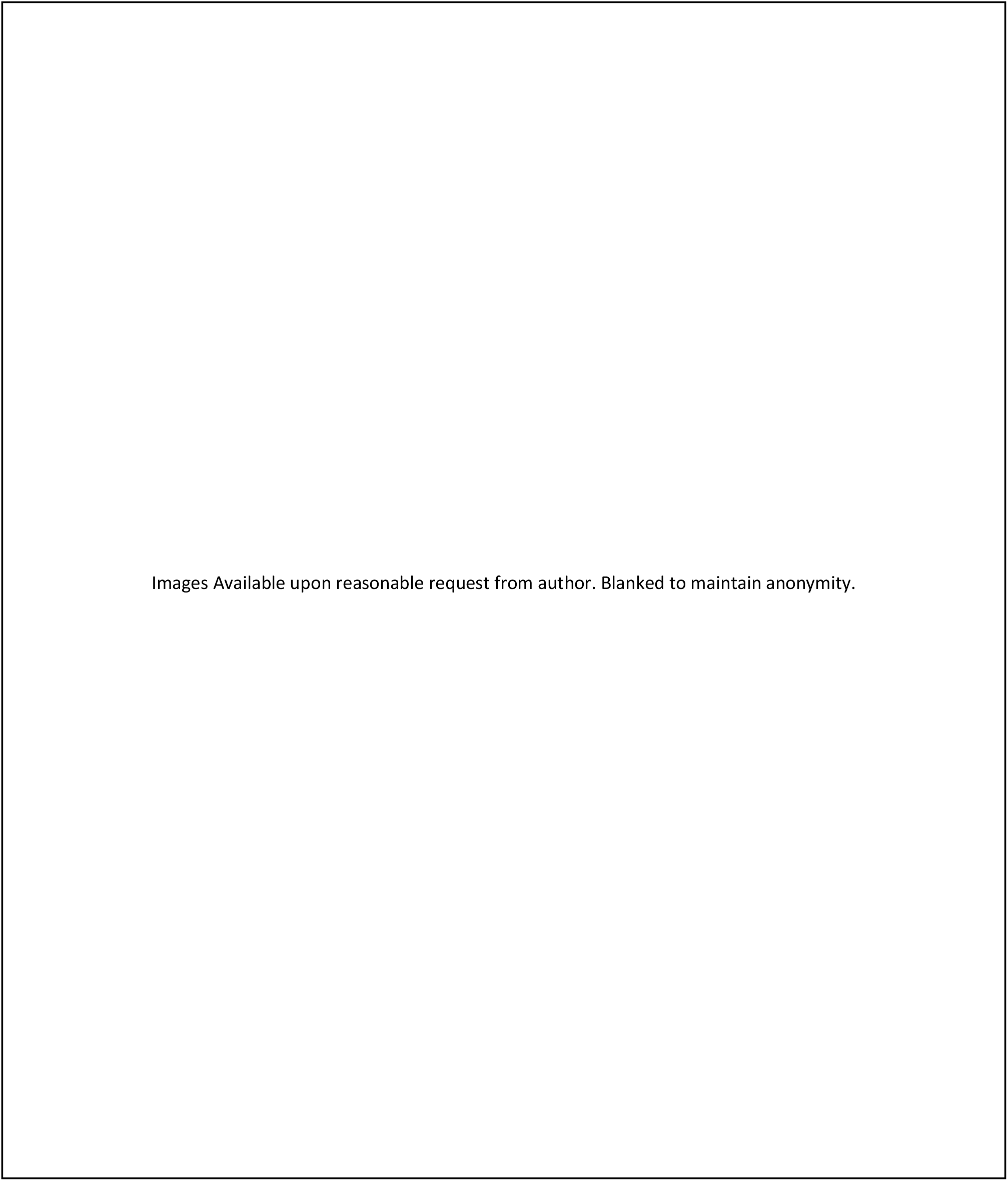
Example of translation in biomechanics from IRS to upright running in Kinogram analysis. Note – Analysis of increased external rotation observed in right limb in IRS and Kinogram (on toe off) at points B compared to left limb at points A.

### Limitations

Despite the injury surveillance data originally being planned across a full season, it is worth noting data collection was reduced to 9 months due to the COVID-19 pandemic in March 2020 with approximately twenty five percent of the season’s games remaining. Injury surveillance data is missing the last 3 months of the season when injuries may be most prevalent, possibly limiting true findings.[3] As is also common in literature of this type, further limitation is present on the information quality of biomechanics as a single point in time (preseason IRS testing) is compared to PCI occurring at multiple points through the season, some months apart.

Additionally, the IRS quantifies an individual score for each participant based on a subjective analysis of movement which will always be open to interpretation as with any descriptive method of movement analysis.[40] This enforces the need for careful undertaking of the methodology in any replication of this study. A new test should demonstrate appropriate levels of reliability and validity and whilst the methodology gives clear direction in performing the IRS system with good pilot levels of IRR, this is a novel test and there is arguably room for development. Further studies could improve the reliability and strength of the IRS in completing a larger IRR analysis. In addition, whilst the inclusion criteria required individuals to be injury free and well rested at the time of IRS testing, no adjustment was made for previous injury history. Amongst athletic populations it is difficult to find athletes without previous injury history, particularly at elite level but as one of the largest risk factors to injury this could be considered a further limitation.

### Future Studies

It may be of interest to first compare the IRS against an established injury predictor such as strength assessment.[23] Any correlation between an established injury risk predictor and IRS system would further strengthen its use as an assessment tool. Performing PHE and PKF is a key component of human movement, particularly in running [16, 27, 41-42] and although the correlation in prone testing to upright running positions has been discussed, formal comparison is required. An upright testing position may provide greater functional comparison although may present difficulties when standardising test position.

## CONCLUSION

The results demonstrate good significant correlation between the novel IRS system compared to PCI within in an athletic population. This study adds evidence that demonstrates that an increased IRS has influence on an individual’s PCI risk and provides guidance for a uniformed assessment tool in assessing this risk. Establishing individual injury risk scores for athletes in this way may aid in preventing and managing PCI that lead to time loss in training and matches. The IRS system can be utilised easily within musculoskeletal settings requiring minimal equipment, making it easily applicable around the constraints of performance-medical environments.

## Supporting information

Line 244

## Data Availability

Deidentified participant data may be available upon reasonable request.

## Acknowledgements

We would like to thank the players of Gloucester Rugby and the performance medical staff who assisted with this study particularly Eoin Power, Steph McNally and Steve Whyte who assisted with injury surveillance data, additionally to Aaron Walters and Luke Croughton who assisted with the inter-rater reliability pilot, and Tom Reynolds who facilitated the video analysis. I (RH) would also like to acknowledge Tony Denton a previous colleague and physiotherapist who originally introduced me to pelvic mechanics and provoked the initial thoughts surrounding this novel screening method, some years ago.

## STATEMENTS

### Contributors

Conception and design: all authors. Acquisition of data: RH. Analysis and interpretation: all authors. Drafting, critical revision and final approval: all authors. Statistical analysis, collection and assembly of the data: RH. Agreement to be accountable for all aspects of the work in ensuring that questions related to the accuracy or integrity of any part of the work are appropriately investigated and resolved: all authors.

### Patient Consent

Informed consent was gained from all participants.

## REFERENCES

1. Mendiguchia J, Alentorn-Geli E, Brughelli, M. Hamstring strain injuries: are we heading in the right direction? British Journal of Sports Medicine 2012;46:81–5.

2. Schuermans J, Danneels L, Van Tiggelen DE, et al. Proximal neuromuscular control protects against hamstring injury in male football players: a prospective study with EMG time-series analysis during maximal sprinting. British Journal of Sports Medicine 2017;51:383–4.

3. Brooks JHM, Kemp SPT. Injury-prevention priorities according to playing position in professional rugby union players. British Journal of Sports Medicine 2011;45:765–75.

4. Askling CM, Tengvar M, Thorstensson A. Acute hamstring injuries in Swedish elite football: a prospective randomised controlled clinical trial comparing two rehabilitation protocols. British Journal of Sports Medicine 2013;47:953–59.

5. Bramah C, Preece SJ, Gill N, et al. Is There a Pathological Gait Associated With Common Soft Tissue Running Injuries? The American Jounral of Sports Medicine, 2018;46:3023–31.

6. Brooks JHM, Fuller CW, Reddin DB. Incidence, risk, and prevention of hamstring muscle injuries in professional rugby union. American Journal of Sports Medicine 2006;34:1297–306.

7. Folland JP, Allen SJ, Black MI, et al. Running technique is an important component of running economy and performance. Medicine and science in sports and exercise 2017;49:1411–2.

8. Mendiguchia J, Edouard P, Samozino P, Brughelli M, et al. Field monitoring of sprinting power-force-velocity profile before, during and after hamstring injury: two case reports. Journal of Sports Sciences, 2016;34:535–41.

9. Gre KG, Millet GP. Conceptual Framework for Strengthening Exercises to Prevent Hamstring Strains. Sports Medicine, 2013;43:1207–15.

10. Opar DA, Williams MD, Timmins, RG, et al. Rate of Torque and Electromyographic Development During Anticipated Eccentric Contraction Is Lower in Previously Strained Hamstrings. American Journal of Sports Medicine, 2013;41:116–25.

11. Ekstrand J, Hagglund M., Walden, M. Injury incidence and injury patterns in professional football: the UEFA injury study. British Journal of Sports Medicine, 2011;45:553–8.

12. Elliott M, Zarins B, Powell JW, et al. Hamstring Muscle Strains in Professional Football Players A 10-Year Review. American Journal of Sports Medicine, 2011;39:843–50.

13. Malliaropoulos N, Papacostas E, Kiritsi O, et al. Posterior Thigh Muscle Injuries in Elite Track and Field Athletes. American Journal of Sports Medicine, 2010;38:1813–9.

14. Lategan L, Gouveia CP. Prevention Of Hamstring Injuries In Sport: A Systematic Review. South African Journal for Research in Sport Physical Education and Recreation, 2018;40:55–69.

15. Duhig S, Shield AJ, Opar D, et al. Effect of high-speed running on hamstring strain injury risk. British Journal of Sports Medicine, 2016;50:1536–40.

16. Bramah C, Preece SJ, Gill N., et al. Is There a Pathological Gait Associated With Common Soft Tissue Running Injuries? The American Jounral of Sports Medicine, 2018;46:3023–31.

17. Van der Horst N, Smits DW, Petersen J, et al. The Preventive Effect of the Nordic Hamstring Exercise on Hamstring Injuries in Amateur Soccer Players: A Randomized Controlled Trial. American Journal of Sports Medicine, 2015;43:1316–23.

18. Malone S, Roe M, Doran DA, et al. High chronic training loads and exposure to bouts of maximal velocity running reduce injury risk in elite Gaelic football. Journal of science and medicine in sport, 2017;20:250–4.

19. Huijing PA. Muscular force transmission necessitates a multilevel integrative approach to the analysis of function of skeletal muscle. Exercise and Sport Sciences Reviews, 2003;31:167–75.

20. Schuermans J, Van Tiggelen D, Palmans T, et al. Deviating running kinematics and hamstring injury susceptibility in male soccer players: Cause or consequence?. Gait & posture, 2017;57:270–7.

21. Schuermans J, Van Tiggelen D, Danneels L, et al. Biceps femoris and semitendinosus—teammates or competitors? New insights into hamstring injury mechanisms in male football players: a muscle functional MRI study. Br J Sports Med, 2014;48:1599–606.

22. Bourne MN, Opar DA, Williams MD, et al. Eccentric Knee Flexor Strength and Risk of Hamstring Injuries in Rugby Union: A Prospective Study. American Journal of Sports Medicine, 2015;43:2663–70.

23. Brockett CL, Morgan DL, Proske U. Human hamstring muscles adapt to eccentric exercise by changing optimum length. Medicine and Science in Sports and Exercise, 2001;33:783–90.

24. Petersen J, Thorborg K, Nielsen MB, Budtz-Jorgensen E, et al. Preventive Effect of Eccentric Training on Acute Hamstring Injuries in Men’s Soccer A Cluster-Randomized Controlled Trial. American Journal of Sports Medicine, 2011;39:2296–303.

25. Brukner P. Hamstring injuries: prevention and treatment-an update. British Journal of Sports Medicine, 2015;49:21–6.

26. Mendiguchia J, Gonzalez De la Flor A, Mendez-Villanueva A, et al. Training-induced changes in anterior pelvic tilt: potential implications for hamstring strain injuries management. Journal of Sports Sciences, 2020;38:1–8.

27. Schuermans J, Van Tiggelen D, Witvrouw E. Prone Hip Extension Muscle Recruitment is Associated with Hamstring Injury Risk in Amateur Soccer. International Journal of Sports Medicine, 2017;38:696–706.

28. Pollock N, James SLJ, Lee JC, et al. British athletics muscle injury classification: a new grading system. British Journal of Sports Medicine, 2014;48:1347–9.

29. McHugh ML. Interrater reliability: the kappa statistic. Biochemia medica: Biochemia medica, 2012;22:276–82.

30. Tortora G, Derrickson B. Principles of anatomy and physiology. 15^th^ ed. United States of America: John Wiley & Sons 2016.

31. IBM. IBM Statistical Package for the Social Sciences (SPSS). Version 25. [For Windows 64]. Portsmouth: IBM 2020.

32. Nacsport. New Assistant for Coach Sport, S.L. [For Windows 64]. United Kingdom: Nacsport 2020.

33. Munro A, Herrington L. The effect of videotape augmented feedback on drop jump landing strategy: implications for anterior cruciate ligament and patellofemoral joint injury prevention. The Knee, 2014;21:891–5.

34. Sado N, Yoshioka S, Fukashiro S. The three-dimensional kinetic behaviour of the pelvic rotation in maximal sprint running. Sports biomechanics, 2017;16:258–71.

35. Pipkin A, Kotecki K, Hetzel S, et al. Reliability of a qualitative video analysis for running. journal of orthopaedic & sports physical therapy, 2016;46:556–61.

36. Van den Tillaar R, Solheim JAB, Bencke J. Comparison of hamstring muscle activation during high-speed running and various hamstring strengthening exercises. International journal of sports physical therapy, 2017;12:717–8.

37. Dingenen B, Barton C, Janssen T, et al. Test-retest reliability of two-dimensional video analysis during running. Physical Therapy in Sport, 2018;33:40–7.

38. Schober P, Boer C, Schwarte. Correlation coefficients: appropriate use and interpretation. Anesthesia & Analgesia, 2018;126:1763–8.

39. Bahr R, Clarsen, B, Derman, W, et al. International Olympic Committee consensus statement: methods for recording and reporting of epidemiological data on injury and illness in sport 2020 (including STROBE Extension for Sport Injury and Illness Surveillance (STROBE-SIIS)). British Journal of Sports Medicine, 2020;54:372–389.

40. McMillan S, Pfaff D. The ALTIS Kinogram Method [Online]. United States of America: ALTIS. Available from https://simplifaster.com/articles/altis-kinogram-method [Accessed 05 December 2020].

41. Herrington L, Munro A. A preliminary investigation to establish the criterion validity of a qualitative scoring system of limb alignment during single-leg squat and landing. Jounral of Exercise and Sports Orthopaedics, 2014;1:1–6.

42. Welling W, Benjaminse A, Lemmink K, et al. Passing return to sports tests after ACL reconstruction is associated with greater likelihood for return to sport but fail to identify second injury risk. The knee, 2020;27:949–57.

